# Improved Performance of ChatGPT-4 on the OKAP Exam: A Comparative Study with ChatGPT-3.5

**DOI:** 10.1101/2023.04.03.23287957

**Authors:** Sean Teebagy, Lauren Colwell, Emma Wood, Antonio Yaghy, Misha Faustina

## Abstract

This study aims to evaluate the performance of ChatGPT-4, an advanced Artificial Intelligence (AI) language model, on the Ophthalmology Knowledge Assessment Program (OKAP) examination compared to its predecessor, ChatGPT-3.5. Both models were tested on 180 OKAP practice questions covering various ophthalmology subject categories. Results showed that ChatGPT-4 significantly outperformed ChatGPT-3.5 (81% vs. 57%; p<0.001), indicating improvements in medical knowledge assessment. The superior performance of ChatGPT-4 suggests potential applicability in ophthalmologic education and clinical decision support systems. Future research should focus on refining AI models, ensuring a balanced representation of fundamental and specialized knowledge, and determining the optimal method of integrating AI into medical education and practice.

## Introduction

The rapid development of artificial intelligence (AI) and natural language processing (NLP) has opened up new possibilities in various domains, including healthcare, education, and research. The application of these foundation models in medicine has been an area of interest, with attempts to have machines take medical qualifying exams. For example, in 2017, news reports claimed that a Chinese AI model called Xiaoyi, which was trained on 2 million medical records and 400,000 articles, was able to pass the Chinese medical licensing exam with a score of 456^1^. More recently, an AI model passed two sets of the UK Royal College of Radiology exam with an overall accuracy of 79.5% compared to 26 radiologists who passed with 84.8% accuracy^2^. The PaLM large language model was recently tested on the United States Medical Licensing Examination (USMLE) and other medical question-answering challenges, including consumer health questions. The results showed a significant improvement over previous AI models, with the PaLM model achieving 67.6% accuracy^3^. OpenAI’s GPT (Generative Pre-trained Transformer) series consistently demonstrates improved language understanding and knowledge representation with each successive iteration. The latest version, ChatGPT-4, has been reported to have superior performance compared to its predecessors^4^. This study aims to evaluate the performance of ChatGPT-4 on the Ophthalmology Knowledge Assessment Program (OKAP) examination compared to ChatGPT-3.5 to determine the potential applicability of this AI model in medical education and clinical practice.

## Methods

The performance of ChatGPT-4 was compared to ChatGPT-3.5 on OKAP practice questions published by the American Academy of Ophthalmology (AAO) under the Basic Clinic and Science Course (BCSC) to evaluate the effectiveness of popular language models in ophthalmologic knowledge^5^. The OKAP exam is an annual, multiple-choice examination administered to ophthalmology residents in the United States, designed to assess their knowledge in various ophthalmology subspecialties. The BCSC, sponsored by the AAO, is a series of OKAP practice questions designed to help resident physicians prepare for the exam.

ChatGPT-3.5 and 4 were provided with the same 180 questions from the BCSC question bank. These questions covered the following ophthalmologic subcategories, as defined by the AAO: Cornea, Neurology, Retina, Optics, Glaucoma, Cataract, Oculoplastics, Fundamentals, Pathology, Pediatrics, Refractory, and Uveitis. ChatGPT-3.5 was queried on December 28th and 29th, 2022, and ChatGPT-4 was queried on March 15th and 16th, 2023. Questions with images in the prompt were removed from the analysis because at the time of querying, ChatGPT could not process images. This resulted in 167 questions being analyzed. Each model was instructed to “select the best answer option and explain why this option was chosen,” followed by each question. If the algorithm did not select an answer option, a second request was used, “please select the best answer option and explain why that option was selected.” The percentage of questions correctly answered was then evaluated according to the answer key provided.

Statistical analysis was performed using SPSS Statistics Software (version 21, SPSS Inc., Chicago, IL, USA). A comparison between the performance of both versions was performed using the Chi-square test. A *p<*0.05 was considered statistically significant.

## Results

ChatGPT-4 performed significantly better than ChatGPT-3.5 (81% vs. 57%; *p*< 0.001) on the 167 OKAP sample questions answered by both models. When comparing each category individually, the performance of ChatGPT-4 was superior to that of ChatGPT-3.5 for all categories other than ‘Fundamentals’ (Graph 1); however, there was not a significant difference due to the small number of questions from each section (Table 1).

**Graph 1:**
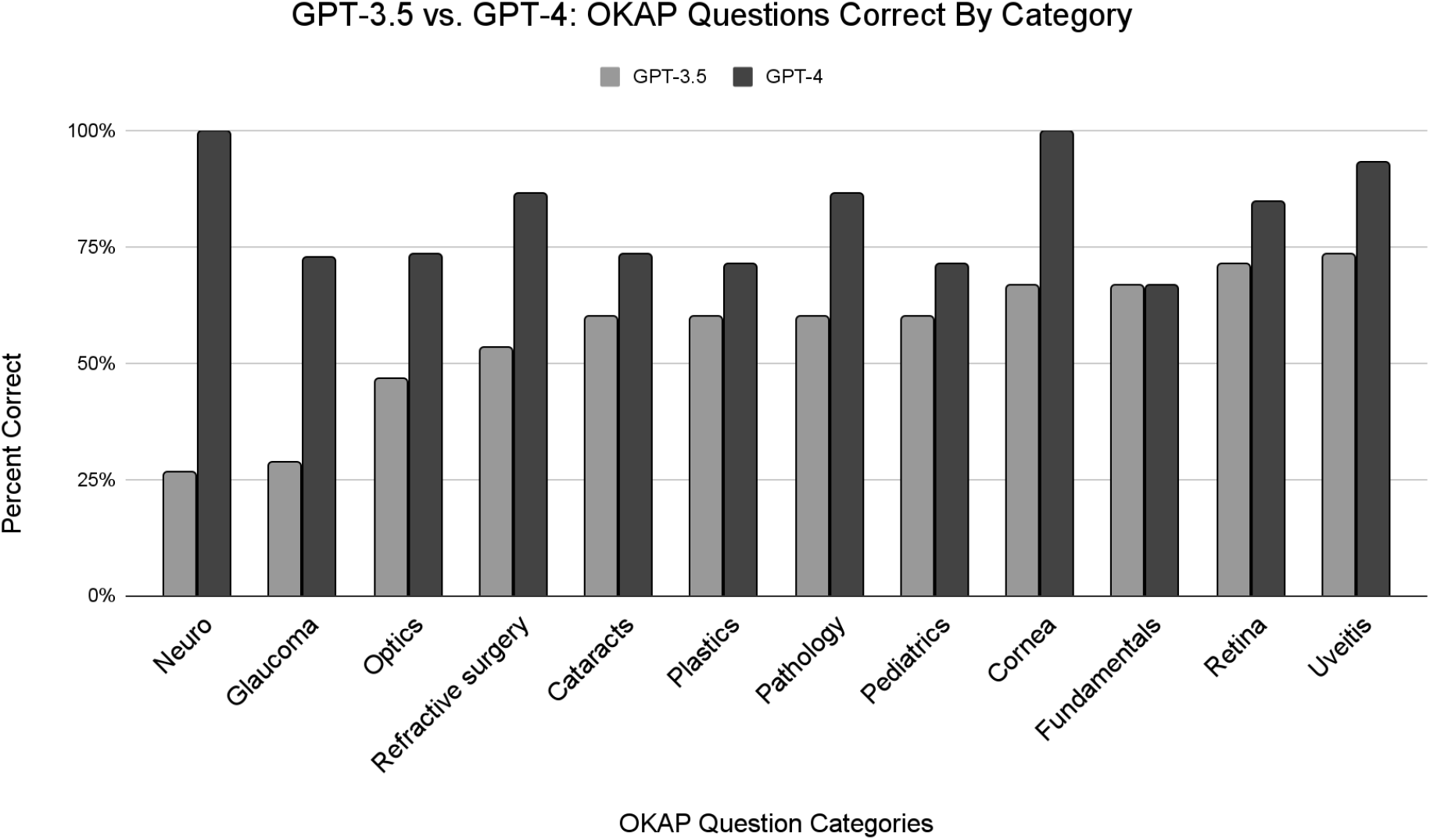
Comparing the performance of ChatGPT-3.5 to ChatGPT-4 on each category of question from the OKAP exam.

**Table 1:** Frequency of correct and incorrect answers by category of OKAP question.

|  | ChatGPT-3.5 |  | ChatGPT-4 |  | Total Questions |
| --- | --- | --- | --- | --- | --- |
|  | Correct answers | Incorrect answers | Correct answers | Incorrect answers |  |
| Cornea | 8 | 4 | 12 | 0 | 12 |
| Neuro | 4 | 9 | 13 | 0 | 13 |
| Retina | 9 | 4 | 11 | 2 | 13 |
| Optics | 7 | 8 | 11 | 4 | 15 |
| Glaucoma | 3 | 8 | 8 | 3 | 11 |
| Cataracts | 9 | 6 | 11 | 4 | 15 |
| Plastics | 9 | 5 | 10 | 4 | 14 |
| Fundamentals | 10 | 5 | 10 | 5 | 15 |
| Pathology | 9 | 6 | 13 | 2 | 15 |
| Pediatrics | 8 | 6 | 10 | 4 | 14 |
| Refractive surgery | 8 | 7 | 13 | 2 | 15 |
| Uveitis | 11 | 4 | 14 | 1 | 15 |
| **Total** | **95** | **72** | **136** | **31** | **167** |

## Discussion

ChatGPT-4 scored significantly higher on the OKAP examination than ChatGPT-3.5. This finding supports the hypothesis that the enhancements made in the ChatGPT-4 model, including architecture improvements, expanded training dataset, which included a more diverse and up-to-date dataset, as well as refined fine-tuning processes, contribute to its superior performance in medical knowledge assessment^4^. The superior performance of ChatGPT-4 has several implications for medical education and AI application in the healthcare sector.

Primarily, ChatGPT-4 provides ophthalmologists with rapid access to a vast amount of medical knowledge that will continue to update and presumably improve with each new version. With a score of 80% correct, ChatGPT-4 scored slightly above the average ophthalmology resident on BCSC questions^6^. It is reasonable to suggest that ChatGPT-4’s improved understanding of medical concepts and reasoning could be leveraged in clinical decision support systems, providing residents with relevant information quickly to aid their decision-making processes. However, the need for improvement in fundamental knowledge questions is necessary because when ChatGPT answers a question incorrectly, it generates text indicating why another answer is correct even though that is not the correct answer. This could be detrimental to learning and could negatively affect both residents and patients if applied to a clinical setting.

Our theory is that ChatGPT-4 did not improve in its performance on questions pertaining to fundamental ophthalmology knowledge because fundamental knowledge represents essential and established information, which inherently would not be as frequently updated in recent literature and databases compared to highly nuanced or specialized topics. Consequently, the model may not have frequently encountered novel data about these fundamental concepts during its updated training. To address this issue, it is crucial to ensure a balanced and comprehensive representation of fundamental and specialized ophthalmology knowledge in the training dataset and to invest in refining the model’s understanding of abstract and general concepts.

Nevertheless, it is essential to recognize the limitations of our study. The models were assessed using multiple-choice questions, which may not fully capture the intricacies of real-world clinical situations. Despite these limitations, the study provides valuable insights into the potential use of AI models in medical education and healthcare. The significant improvement of ChatGPT-4 over ChatGPT-3.5 in the OKAP examination serves as an indicator of the rapid advancement of AI capabilities in the medical domain. However, it is crucial to approach the integration of AI into medical practice with caution, as ethical considerations, potential biases, and the importance of human interaction in patient care must be thoroughly considered.

ChatGPT could be used to complement traditional learning methods and not as a replacement for human instruction, mentorship, or care delivery. Integrating AI models in medical education and practice have the risk of potential biases, unknown ethical approaches, and the loss of human interaction in patient care^7,8^. Future research should focus on the applicability of ChatGPT-4 with particular attention focusing on the slower response rate of more advanced ChatGPT models ^4^. Additionally, further investigation should be conducted to determine the optimal method of integrating AI models into ophthalmology education and clinical practice, ensuring that these tools are used effectively and ethically.

In conclusion, our study reveals that ChatGPT-4 significantly outperforms ChatGPT-3.5 on the OKAP examination, indicating the potential for enhanced AI models to support medical education and practice. As AI continues to advance, it is essential for the medical community to remain engaged with these developments, ensuring that the potential benefits of AI are maximized while minimizing the risks associated with its implementation in the healthcare sector.

## Data Availability

All data produced in the present study are available upon reasonable request to the authors

